# Resting-State Functional Connectivity and Cognitive Performance in Long COVID: A Randomized Controlled Trial

**DOI:** 10.64898/2026.10.05.26364531

**Authors:** David O’Connor, Eric Watson, David L. Reich, Fernando Carnavali, Megan Mcveety, Pranav Gwalani, Juan P. Wisnivesky, Jacqueline H. Becker

## Abstract

Cognitive impairment is a prevalent, disabling manifestation of Long COVID, but whether cognitive rehabilitation alters neural network organization is unknown. We examined whether a 12-week cognitive rehabilitation intervention modified resting-state connectivity within the central-executive (CEN), default-mode (DMN), and salience (SN) networks, and whether connectivity changes associated with cognitive performance. Participants were adults with prior SARS-CoV-2 infection, persistent Long COVID symptoms (≥3 months), and objectively confirmed mild-to-moderate cognitive impairment. Of 58 participants randomized, 39 (mean age 46.4; 69% women) completed neuroimaging (19 intervention, 20 control). The 12-week intervention combined virtual cognitive rehabilitation and small-group and individual sessions with computerized cognitive training. No significant group, time, or group-by-time interaction effects were observed for within- or between-network connectivity (all P > 0.22). Exploratory analyses identified nominal associations (P < 0.05) between connectivity changes and cognition; improved processing speed associated with increased CEN-DMN connectivity, greater SN-DMN connectivity associated with worse performance across processing speed, working memory, and executive function. None survived correction for multiple comparisons. Cognitive rehabilitation was not associated with detectable changes in resting-state network connectivity in long COVID–related cognitive impairment. SN-DMN connectivity may represent a marker of reduced network segregation underlying cognitive dysfunction, warranting study in larger, multimodal trials. Trial registration: NCT05498493.

## 1. Introduction

Post-acute sequelae of SARS-CoV-2 (PASC), commonly referred to as long COVID, affects an estimated 10-30% of adult individuals following acute infection and represents a significant public health concern [1], [2]. Among the constellation of persistent symptoms, cognitive dysfunction (“brain fog”) is one of the most prevalent and disabling. Post-COVID cognitive dysfunction spans multiple domains, though executive dysfunction appears most prominent [3], [4] and may persist over time [5].

The neurobiological mechanisms underlying these symptoms remain incompletely understood, but converging evidence implicates disruption of large-scale brain networks. Resting-state functional magnetic resonance imaging (rs-fMRI) studies have identified altered functional connectivity in individuals with post-COVID cognitive symptoms [6], [7], [8], [9]. Functional connectivity reflects the temporal correlation of neural activity across distributed brain regions and indexes the integrity of large-scale networks supporting cognition [10], [11], [12]. Three networks are especially relevant: the Central Executive Network (CEN), supporting working memory and cognitive control; the Default Mode Network (DMN), involved in self-referential and memory processes; and the Salience Network (SN), which mediates switching between internally and externally directed attention [6], [8]. Disruptions in the balance among these networks has been linked to cognitive dysfunction across neurological and psychiatric conditions [13], [14], [15].

Cross-sectional studies in long COVID have reported reduced connectivity within and among these canonical networks, including reduced connectivity within the CEN and DMN, altered integration between networks, and associations with cognitive performance [6], [8]. However, this literature is limited by small sample sizes, heterogeneity, and a lack of longitudinal data examining whether these alterations are modifiable. In other conditions (e.g., multiple sclerosis), cognitive rehabilitation has been associated with changes in functional connectivity and brain activation that correlates with cognitive improvements [16], [17]. Whether similar neural plasticity occurs in long COVID is unknown.

We recently conducted a pilot randomized controlled trial of a 12-week cognitive rehabilitation program for individuals with long COVID and cognitive dysfunction. In our initial report, we demonstrated the intervention was feasible and associated with modest improvements in cognitive patient-reported outcomes compared with an attention-matched control condition [18]. However, the neural mechanisms underlying these changes remain unclear.

The present study examined whether cognitive rehabilitation modifies functional brain connectivity in long COVID. Using seed-based analyses of the CEN, DMN, and SN, we evaluated changes within- and between-network connectivity and their associations with cognitive performance. We hypothesized that the intervention would enhance CEN and DMN connectivity and that connectivity changes would correlate with cognitive improvement.

## 2. Methods

### 2.1 Participants and Study Design

This parallel-group randomized controlled trial recruited participants from the Mount Sinai Health System (New York City) between September 2022 and August 2024. Eligible participants were adults (≥18 years) with documented SARS-CoV-2 infection, persistent long COVID symptoms (≥3 months), and objectively confirmed mild-to-moderate cognitive impairment (≥1.5 standard deviations below normative means in executive function and one additional domain) [19], [20]. Exclusion criteria included pre-existing cognitive impairment, major neurological conditions, significant traumatic brain injury, active substance use disorder, and unstable medical or psychiatric illness. Individuals who were concurrently engaged in other cognitive remediation programs or regular meditative practices were also ineligible. The study protocol received approval from the Icahn School of Medicine at Mount Sinai Institutional Review Board, and all participants provided written informed consent. Full details of the randomization procedure, allocation concealment, blinding, and sample size determination for the parent trial are reported in Becker et al. [18] and the trial protocol is available at clinicaltrials.gov (NCT05498493). A participant flow chart is shown in **Figure 1**. This report adheres to CONSORT guidelines for randomized trials. No adverse events related to the intervention or imaging procedures were reported during the study period.

**Figure 1.**
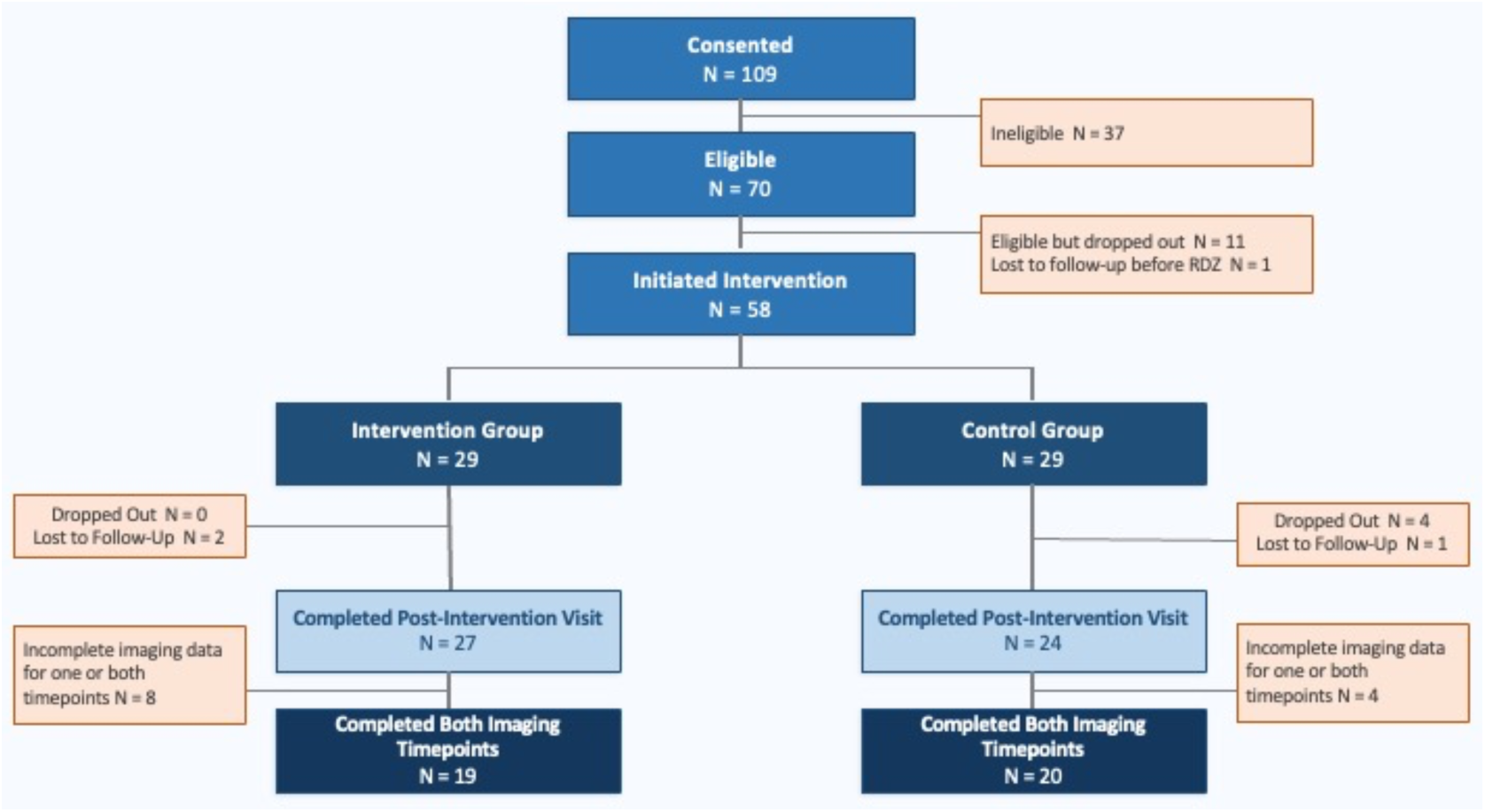
Participant flow diagram. Enrollment, randomization, and retention for the parent randomized controlled trial and neuroimaging subsample. Of 58 participants who initiated the intervention, 39 completed resting-state fMRI at both timepoints and were included in neuroimaging analyses (19 intervention, 20 control). Participants excluded from neuroimaging analyses had incomplete imaging data at one or both timepoints due to motion or image quality.

### 2.2 Cognitive Assessments

Participants completed standardized neuropsychological testing at baseline (pre-intervention) and within two weeks post-intervention. Executive function (EF) was assessed using three complementary EF measures: Trail Making Test (TMT)- Part B (cognitive flexibility and set-shifting) [21], the Wisconsin Card Sorting Test–64 card version (WCST; abstract reasoning and novel problem solving), and the Delis–Kaplan Executive Function System (D-KEFS) Color– Word Interference Test (inhibitory control). Attention and working memory were assessed using Number Span Forward (attention) and Number Span Backward (working memory) [22]. Processing speed was measured using TMT-Part A and the D-KEFS Color Naming and Word Reading. Verbal fluency was assessed using the D-KEFS subtest (phonemic and semantic). Verbal learning and memory were evaluated using the Hopkins Verbal Learning Test–Revised (HVLT-R; immediate, delayed recall, and recognition) [23].

Perceived cognitive functioning was assessed using the Cognitive Failures Questionnaire (CFQ), capturing patient-reported cognitive difficulties. Alternate test forms were used when available to minimize practice effects. Raw scores were transformed into demographically corrected *z*-scores using age-, sex-, and education-adjusted normative data [24].

### 2.3 Neuroimaging Methods

#### 2.3.1 Data Acquisition

All imaging was performed on a Siemens MAGNETOM 3T scanner using a 32-channel head coil. High-resolution structural images were acquired using a sagittal T1-weighted MPRAGE sequence with the following parameters: repetition time (TR) = 2400 ms, echo time (TE) = 2.07 ms, inversion time (TI) = 1000 ms, flip angle = 9°, field of view (FOV) = 256 mm × 256 mm, matrix size = 320 × 320, 224 sagittal slices, and isotropic voxel size = 0.8 mm³. Parallel imaging was applied using GRAPPA with an acceleration factor of 2.Resting-state functional MRI (rs-fMRI) data were collected using a multiband echo-planar imaging (EPI) sequence with the following parameters: TR = 1000 ms, TE = 35.00 ms, flip angle = 60°, FOV = 228 mm × 228 mm, matrix = 108 × 108, 70 contiguous axial slices acquired in interleaved order with a multiband acceleration factor of 7, and isotropic voxel size = 2.1 mm³. Each rs-fMRI run lasted 5 minutes and 33 seconds and consisted of 322 volumes. A total of three rs-fMRI runs were collected per participant. Fat saturation was applied; no online motion correction or spatial filtering was performed.

#### 2.3.2 MRI Preprocessing

All MRI data were preprocessed using fMRIPrep 23.2.1 [25], which integrates tools from ANTs, FSL, FreeSurfer, and other packages within a standardized workflow. Internal processing steps relied on Nilearn 0.10.4 [26].

##### 2.3.2.1 Structural MRI Preprocessing

Each subject contributed two high-resolution T1-weighted (T1w) anatomical images. Intensity non-uniformity was corrected using ANTs, and the images were skull-stripped using a template-based approach. Tissue segmentation into gray matter (GM), white matter (WM), and cerebrospinal fluid (CSF) was performed with FSL. A robust average T1w reference image was created by aligning and combining the two anatomical scans. Nonlinear spatial normalization to MNI152NLin2009cAsym was then performed using ANTs. Templates and atlases were accessed via TemplateFlow [27].

##### 2.3.2.2 Functional MRI Preprocessing

Each subject underwent six BOLD fMRI runs across two sessions. Susceptibility-induced distortion was corrected using field maps with opposing phase-encoding directions, using FSL [28]. The corrected BOLD reference was co-registered to the subject’s anatomical reference image using FSL and FreeSurfer [29]. All transformations—including motion correction, distortion correction, co-registration, and normalization to standard space—were combined into a single resampling step to minimize interpolation artifacts.

Nuisance regressors were estimated from the preprocessed BOLD data, including framewise displacement (FD), and global signal fluctuations from CSF, WM, and whole-brain masks. Physiological noise components were extracted using anatomical and temporal variants of the CompCor method [30]. For anatomical CompCor, WM and CSF masks were used to extract noise components after excluding voxels with substantial gray matter partial volume. For temporal CompCor, components were computed from the most variable voxels across the brain.

All confound time series were expanded with temporal derivatives and quadratic terms [31]. Volumes exceeding FD > 0.5 mm or standardized DVARS > 1.5 were flagged as motion outliers. Additional nuisance regressors were estimated from signal fluctuations near the cortical surface using a principal component analysis of peripheral “crown” voxels, following published guidelines [32].

#### 2.3.3 Seed-Based Connectivity Analysis

We defined 19 seed regions of interest (ROIs) representing three major brain networks: the Central Executive Network (CEN; 10 regions), Salience Network (6 regions), and Default Mode Network (DMN; 3 regions). Seed coordinates were based on established network atlases and prior literature and are shown in **Table 1**. CEN seed ROIs were defined using a 5-mm radius sphere centered at coordinates for 10 CEN nodes [33], identified via independent components analysis and included in a publicly available atlas of resting state networks [34]. Seeds for the DMN and Salience networks were similarly generated from Anderson et al. and Nusslock et al. respectively [35], [36].

**Table 1.** MNI coordinates of brain regions used as seeds for the functional network analysis.

| **Network** | **Region** | **Hemisphere** | **MNI Coordinates**<br>**(x, y, z)** |
| --- | --- | --- | --- |
| **CEN** | Inferior parietal lobule | Left | (-42, -63, 46) |
|  | Middle frontal gyrus 1 | Left | (-32, 23, 49) |
|  | Middle frontal gyrus 2 | Left | (-40, 48, -1) |
|  | Middle temporal gyrus | Left | (-59, -42, -12) |
|  | Medial frontal gyrus | Left | (-7, 34, 43) |
|  | Middle frontal gyrus 1 | Right | (38, 26, 42) |
|  | Middle frontal gyrus 2 | Right | (38, 54, 1) |
|  | Inferior parietal lobule | Right | (48, -54, 47) |
|  | Caudate | Right | (13, 2, 14) |
|  | Medial frontal gyrus | Right | (6, 37, 46) |
| **Salience** | Dorsal anterior cingulate cortex (dACC) | Left | (-6, 17, 47) |
|  | Middle frontal gyrus | Left | (-31, 47, 22) |
|  | Anterior insula<sup>1</sup> | Left | (-34, 16, -4) |
|  | Dorsal anterior cingulate cortex (dACC) | Right | (6, 17, 47) |
|  | Middle frontal gyrus | Right | (28, 46, 26) |
|  | Anterior insula<sup>1</sup> | Right | (38, 16, 0) |
| **DMN** | Posterior cingulate cortex (PCC) | Midline | (-8, -56, 26) |
|  | Medial prefrontal cortex (mPFC) | Midline | (-6, 52, -2) |
|  | Temporoparietal junction (TPJ) | Left | (-54, -54, 28) |

<sup>1</sup> Left and right anterior insula coordinates are selected based on peak z-scores from Neurosynth's meta- analysis for "anterior insula" to avoid duplication found in the Nusslock et al. dataset.

For each participant and session, connectivity matrices were computed by extracting average time series from each seed region and calculating Pearson correlations between all region pairs. Correlation values were Fisher *z*-transformed using the arctanh function to normalize distributions. An example illustration is shown in **Figure 2**.

**Figure 2.**
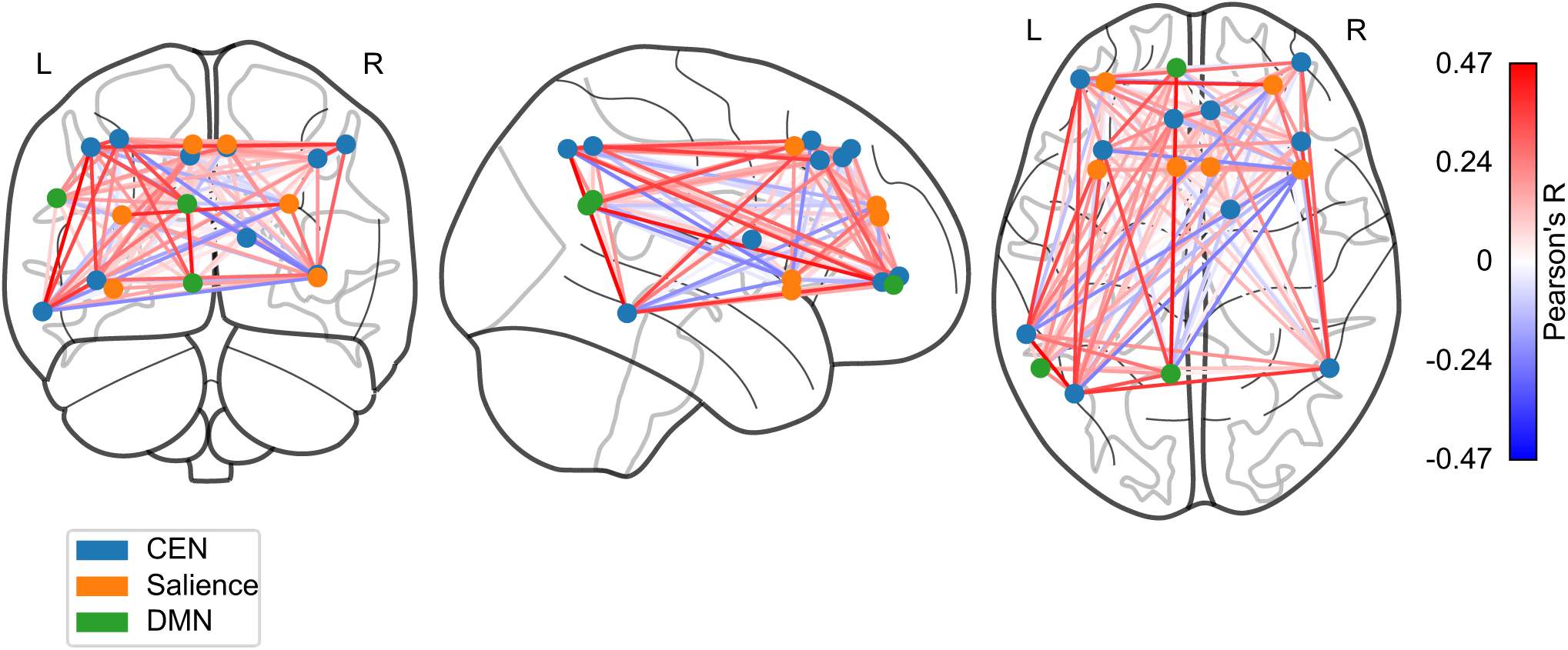
Network visualization of functional connectome in the control group at baseline. Node positions represent anatomical coordinates of brain regions, with node colors indicating network membership: CEN (Central Executive Network, blue), Salience (orange), and DMN (Default Mode Network, green). Edge thickness and color intensity reflect the strength of functional connectivity between regions, with red edges indicating positive correlations and blue edges indicating negative correlations.

#### 2.3.4 Network-Level Connectivity Metrics

We computed several network-level summary measures (**Figure 3**):

1. Overall network mean: Average connectivity across all seed pairs.
2. Within-network connectivity: Mean connectivity within CEN, Salience, and DMN networks separately.
3. Between-network connectivity: Mean connectivity between network pairs (CEN-Salience, CEN-DMN, Salience-DMN)

**Figure 3.**
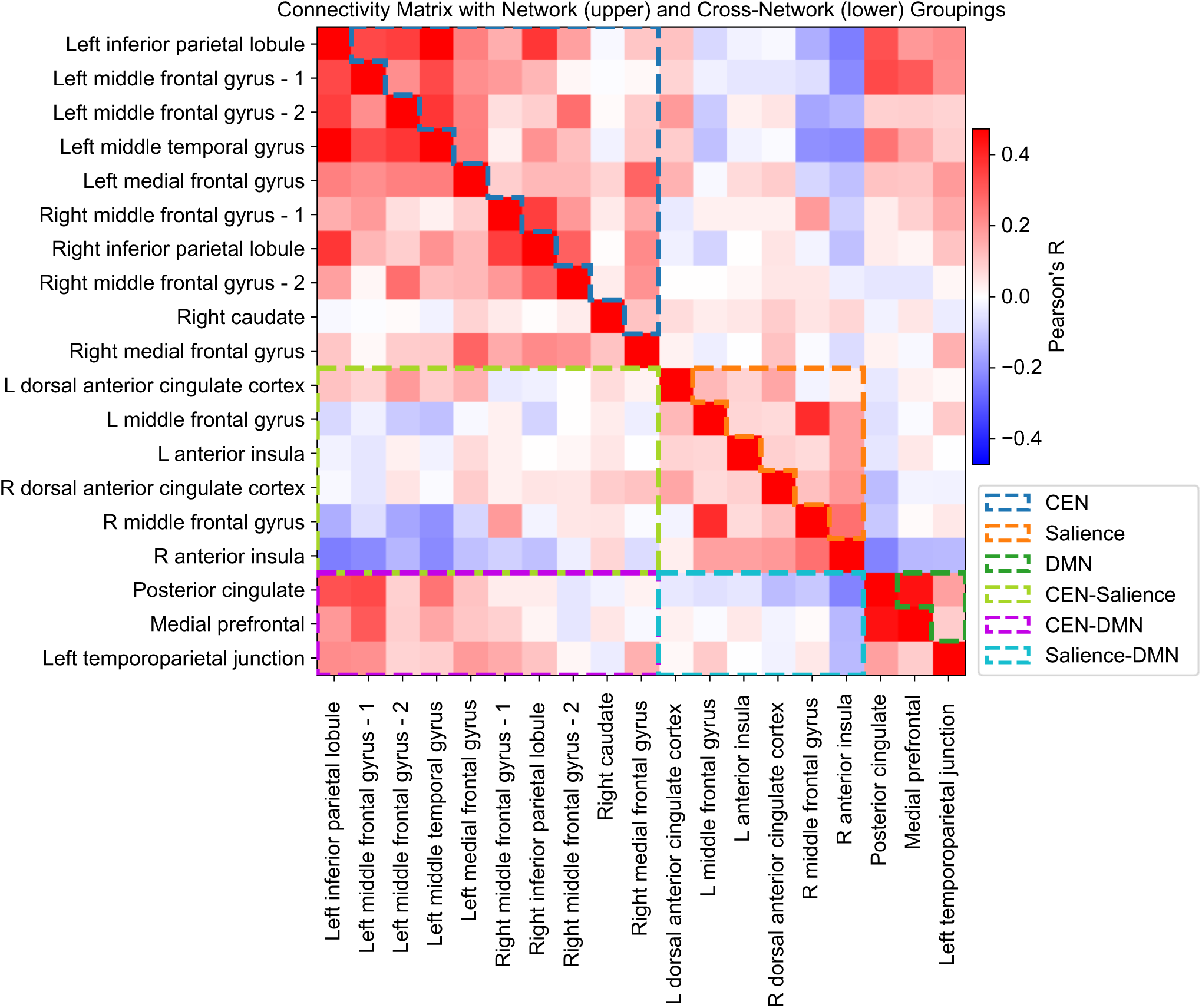
Connectivity matrix visualization showing functional connectivity strength between brain regions in the control group at baseline. The matrix displays correlation coefficients between all pairs of brain regions, with heatmap colors representing connectivity strength (red = positive correlation, blue = negative correlation). Dashed boxes highlight network organization: within-network connectivity is shown in the upper diagonal with CEN (blue), Salience (orange), and DMN (dark green) networks outlined along the diagonal, while cross-network connectivity patterns are displayed in the lower diagonal with CEN-Salience (bright green), CEN-DMN (purple), and Salience-DMN (cyan) interactions.

For each participant, multiple runs within each session were averaged to improve signal reliability. Connectivity matrices for participants with full data at baseline and follow-up sessions were retained for longitudinal analyses.

#### 2.3.5 Quality Control and Confound Regression

Motion quality was assessed using mean framewise displacement. To control for nuisance variables, we regressed out age, gender, and mean FD from all network connectivity measures, retaining residual values for subsequent analyses.

### 2.4 Statistical Analyses

#### 2.4.1 Cross-sectional Associations

We examined associations among network connectivity measures and cognitive performance using ordinary least squares regression. Cognitive variables included standardized scores from tests of Attention (Number Span Forward), Working Memory (Number Span Backward), Executive Functioning (TMT-B, WCST, D-KEFS Inhibition), Language (D-KEFS verbal fluency), Learning & Memory (HVLT-R), Processing Speed (TMT-A, D-KEFS Word Reading/Color Naming), and Subjective Cognitive Complaints (CFQ).

#### 2.4.2 Longitudinal Mixed-Effects Models

For participants with complete data at both timepoints, we implemented linear mixed-effects models with random intercepts to account for within-subject correlation. Models tested:

1. Main effects: Timepoint (baseline vs. follow-up) and intervention group (treatment vs. control)
2. Interaction effects: Timepoint × intervention group interactions

#### 2.4.3 Change Score Analyses

We computed change scores (Session 1 - Session 2) for both connectivity and cognitive measures. Linear regression models examined associations among connectivity changes and cognitive changes, with separate analyses for treatment and control groups.

## 3 Results

Fifty-eight participants enrolled; 39 completed both imaging sessions (mean [SD] age, 46.4 [12.3] years; 69% female). Nineteen participants were in the intervention group and 20 in the control group.

### 3.1 Effects of intervention on connectivity

Linear mixed-effects models did not reveal any statistically significant effects of Timepoint, Intervention, or their interaction on network connectivity metrics (**Supplemental Table 1**). For the overall network mean, neither Timepoint (β = -0.005, 95% CI = [-0.015, 0.005], t = -0.913, *p* = 0.36), Intervention (β = -0.011, 95% CI = [-0.024, 0.002], t = -1.614, *p* = 0.11), nor their interaction (β = 0.003, 95% CI = [-0.011, 0.018], t = 0.436, *p* = 0.66) reached significance. Similarly, no significant effects were observed within the canonical resting state networks; the CEN, salience network, or DMN, all *p*-values > 0.25. Connectivity between network pairs (e.g., CEN-DMN, CEN-salience, and salience-DMN) also showed no significant changes. Boxplots of these data are in **Supplemental Figures 1 and 2**. Overall; these findings suggest that the intervention did not produce detectable changes in within- or between-network functional connectivity.

### 3.2 Brain-behavior associations

Prior work on this dataset showed that while intervention effects on cognitive measures were minimal, timepoint effects were significant, as all participants showed improvement with time across executive function, memory and self-reported cognition [18]. Since intervention effects were also not detectable in the network measures we chose to examine if the changes in cognition were related to changes in network measures. We calculated the timepoint delta for all imaging and non-imaging measures and modelled the pairwise relationship between each imaging metric, and each of the non-imaging measure **(Figure 4**).

**Figure 4.**
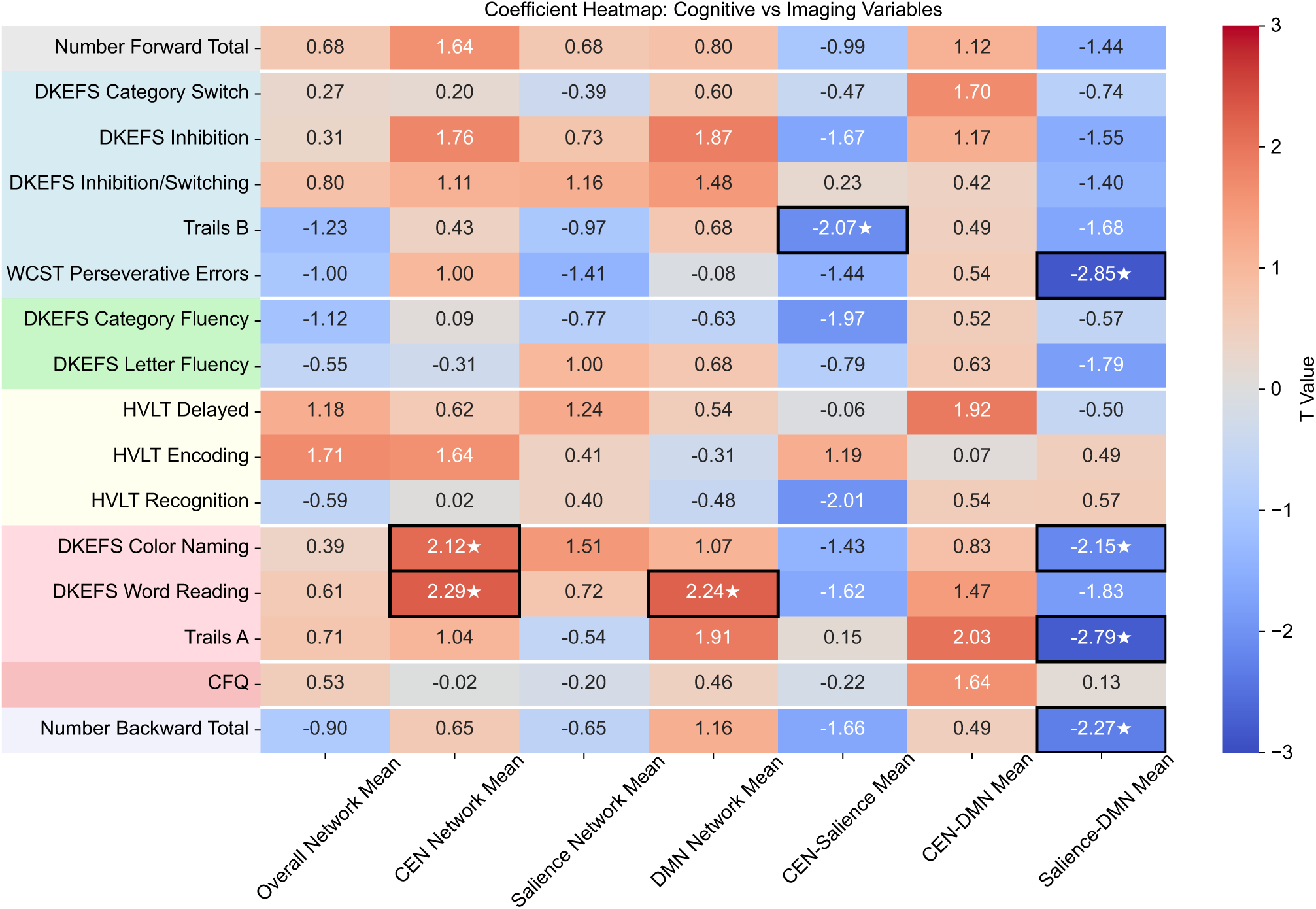
Associations among cognitive performance and brain network connectivity changes. Heatmap showing standardized regression coefficients from linear models examining relationships between cognitive test performance and brain network connectivity measures (controlling for age, head motion, and sex). Rows represent cognitive variables grouped by domain with colored background indicators: Attention (grey; Number Span Forward), Executive Functioning (light blue; Trail Making Test B, Wisconsin Card Sorting Test, D-KEFS Inhibition measures), Language (green; D-KEFS fluency measures), Learning & Memory (light yellow; Hopkins Verbal Learning Test variants), Processing Speed (pink; D-KEFS Word Reading and Color Naming, Trail Making Test A), Subjective Complaints (light red; Cognitive Failures Questionnaire), and Working Memory (lavender; Number Span Backward). Columns represent brain network connectivity measures: overall network connectivity, within-network connectivity (CEN, Salience, DMN), and between-network connectivity (CEN-Salience, CEN-DMN, Salience-DMN). Cells marked with ★ indicate statistically significant associations (p < 0.05), with black borders highlighting significant relationships. Significant associations have not been corrected for multiple comparisons. CEN = Central Executive Network; DMN = Default Mode Network.

Eight nominal associations (p<0.05) were observed. Processing speed measures showed the strongest patterns, with DKEFS Word Reading demonstrating positive associations with both CEN Network connectivity (β = 9.64, 95% CI = [1.09, 18.19], t = 2.29, p = 0.028) and DMN Network connectivity (β = 3.54, 95% CI = [0.33, 6.75], t =2.24, p = 0.032), while DKEFS Color Naming exhibited a positive association with CEN Network connectivity (β = 7.83, 95% CI = [0.35, 15.32], t = 2.12, p = 0.041) and a negative association with Salience-DMN cross-network connectivity (β = -5.6, 95% CI = [-10.88, -0.31], t = -2.15, p = 0.039). Processing speed (Trails A) also showed a negative association with Salience-DMN cross-network connectivity (β = - 9.29, 95% CI = [-16.05, -2.53], t = -2.79, p = 0.008). Working memory performance, measured by Number Backward Total, showed a negative association with Salience-DMN cross-network connectivity (β = -4.67, 95% CI = [-8.83, -0.5], t = -2.27, p = 0.029). Executive functioning showed two significant relationships; Trails B performance demonstrated a negative association with CEN-Salience cross-network connectivity (β = -16.94, 95% CI = [-33.52, -0.36], t = -2.07, p = 0.045), and WCST Perseverative Errors demonstrated a negative association with Salience-DMN cross-network connectivity (β = -5.97, 95% CI = [-10.23, -1.72], t = -2.85, p = 0.007).

This pilot (N = 39) was not powered to detect small-moderate effects in resting-state functional connectivity. For a two-group, two-timepoint mixed-effects design with repeated measures, the minimum detectable standardized effect for the Timepoint × Intervention interaction was in the moderate-large range (80% power to detect Cohen’s *d* ≈ 0.7, assuming α = 0.05). Accordingly, the null findings do not exclude the presence of smaller intervention-related neural changes. These results should therefore be interpreted as preliminary and hypothesis-generating, with larger, adequately powered trials required to determine whether cognitive rehabilitation produces modest alterations in large-scale network organization.

## 4. Discussion

In this randomized trial, cognitive rehabilitation was not associated with measurable changes in resting-state connectivity across major brain networks. However, changes in connectivity were associated with cognitive performance, suggesting that network dynamics remain behaviorally relevant even in the absence of detectable intervention effects. The absence of group-level effects mirror prior behavioral findings [18], in which both groups improved over time. However, while the intervention did not produce detectable changes in large-scale networks, the associations observed suggest that neural changes do accompany cognitive improvement, they simply may not be intervention-specific or require more intensive treatment to manifest as group-level differences.

Several explanations may account for the null findings. First, cognitive rehabilitation may act through mechanisms not captured by static rs-fMRI (e.g., dynamic connectivity or compensatory strategies). The significant improvement in self-reported cognitive functioning in the intervention group, despite modest objective test changes, supports this possibility. Second, intervention dose or duration may have been insufficient to induce detectable network-level changes, suggesting more intensive or prolonged treatment may be required to demonstrate group-level shifts. Third, resting-state measures may lack sensitivity to short-term treatment effects, when compared to behavior specific in-scanner tasks.

Despite null group effects, consistent brain-behavior associations emerged. Processing speed improvements were associated with increased CEN and DMN connectivity, suggesting enhanced efficiency of task-relevant networks. In contrast, increased SN-DMN connectivity was associated with worse performance across multiple domains. This pattern aligns with models emphasizing the importance of network segregation. Excess coupling between the SN and DMN may reflect impaired switching between internally and externally directed attention, contributing to cognitive inefficiency. The CEN encompasses frontoparietal regions supporting controlled, effortful cognition, such that stronger within-CEN connectivity may reflect more efficient recruitment of these regions during processing-speed demands.

Notably, SN-DMN connectivity was the only network metric to show significant associations across multiple cognitive domains, correlating negatively with measures of processing speed (D-KEFS Color Naming, TMT-A), working memory (Number Span Backward), and executive function (WCST Perseverative Errors). Because all cognitive variables were expressed as demographically corrected *z*-scores in which higher values indicate better performance, the uniformly negative direction of these associations indicates that greater Salience-DMN connectivity was consistently associated with poorer cognitive outcomes across domains. Although none of these individual associations would survive correction for multiple comparisons, this directional consistency across four measures spanning three cognitive domains is notable and suggests that SN-DMN connectivity may represent a particularly sensitive marker of network disorganization in this population. The WCST Perseverative Errors finding is of particular mechanistic interest. Perseverative errors reflect a failure of cognitive flexibility or the inability to disengage from a previously reinforced rule in response to negative feedback. The salience network is theorized to mediate precisely this type of behavioral switching by detecting when current strategies are no longer effective and signaling the CEN to update. Elevated SN-DMN coupling may impair the salience network’s capacity to generate clear switching signals, resulting in perseverative behavior. More broadly, this cross-domain pattern aligns with network neuroscience models in which blurring of boundaries between intrinsic networks, particularly between task-negative (DMN) and task-positive systems, is associated with worse cognitive performance, as has been observed in aging and other neurological conditions. These findings, while preliminary and in need of replication, suggest that SN-DMN segregation may be a candidate biomarker worth examining in future, adequately powered studies of cognitive dysfunction in long COVID.

The negative association between CEN-SN connectivity and TMT-B performance warrants particular attention given that TMT-B improvements were virtually identical between groups in the behavioral trial (DID: −0.01). This pattern suggests that while both groups improved equally on this measure, the neural correlates of that improvement involved reduced coupling between the CEN and SN. The SN plays a critical role in detecting behaviorally relevant stimuli and facilitating appropriate network engagement. Tighter coupling with the CEN may introduce interference or reduce functional specialization, impeding performance on tasks requiring cognitive flexibility and set-shifting.

Several limitations should be considered. The sample size, while adequate for the pilot trial’s futility design, may have been underpowered to detect mild-to-moderate intervention effects on resting state connectivity measures. Resting state fMRI typically has smaller effect sizes and more participant motion and drowsiness in scanner than task fMRI. A low demand movie paradigm, such as inscapes [37], could be used to boost power in future studies. fMRI can also be combined with diffusion and structural MRI to conduct multimodal analyses which may have higher test-retest reliability and larger effects sizes [38], [39], [40]. The relatively highly educated nature of our sample may limit generalizability. Future trials with larger samples, longer intervention durations, or multimodal imaging approaches may be better positioned to detect neural mechanisms of treatment response.

In conclusion, this neuroimaging analysis of a pilot randomized controlled trial found no evidence that cognitive rehabilitation altered resting-state functional connectivity in individuals with long COVID-related cognitive impairment, consistent with the absence of significant group differences in objective cognitive outcomes. Nevertheless, changes in processing speed, working memory, and executive function tracked with within-network connectivity in executive and default mode regions, and SN-DMN connectivity emerged as a consistent cross-domain correlate of worse cognitive performance. These findings underscore the complexity of neural mechanisms underlying cognitive dysfunction in long COVID and highlight the need for larger, adequately powered trials to determine whether more intensive interventions can produce both behavioral and neural improvements in this population.

## Supporting information

RCT Protocol

## Data Availability

Data will be shared upon reasonable request.

## Acknowledgments

We thank all participants for contributing their time to this study and for their commitment to this research. This work was completed in accordance with the Helsinki Declaration.

## Data Sharing Statement

Data will be shared upon reasonable request.

## CRediT Author Contributions

- DOC: Formal analysis, Visualization, Writing – original draft, Writing – review and editing
- EW: Project administration, Supervision, Writing – review and editing
- DLR: Funding acquisition, Writing – review and editing
- FC: Project administration, Writing – review and editing
- MM: Project administration, Writing – review and editing
- PG: Formal Analysis, Data Curation
- JPW: Conceptualization, Funding acquisition, Investigation, Methodology, Project administration, Supervision, Writing – review and editing
- JHB: Formal analysis, Conceptualization, Funding acquisition, Investigation, Methodology, Project administration, Supervision, Writing – original draft, Writing – review and editing During the preparation of this work the authors used Claude Sonnet 4.6 in order to edit and summarize text. After using this tool, the authors reviewed and edited the content as needed and take full responsibility for the content of the published article.

