## Supplemental Information for "Resting-State Functional Connectivity and Cognitive Performance in Long COVID: A Randomized Controlled Trial"

**Supplemental Table 1 – Results of mixed effects linear modelling of the effect of intervention and timepoint on the functional network measures.**

| Variable | Category | Beta | 95% CI | T-Value | P-value |
|---|---|---|---|---|---|
| **Overall network mean** | Timepoint | -0.0047 | [-0.015, 0.005] | -0.913 | 0.36 |
| **Overall network mean** | Intervention | -0.0109 | [-0.024, 0.002] | -1.614 | 0.11 |
| **Overall network mean** | Timepoint x Intervention | 0.0032 | [-0.011, 0.018] | 0.436 | 0.66 |
| **CEN** | Timepoint | -0.0108 | [-0.030, 0.009] | -1.084 | 0.28 |
| **CEN** | Intervention | -0.0005 | [-0.036, 0.035] | -0.026 | 0.98 |
| **CEN** | Timepoint x Intervention | 0.0079 | [-0.020, 0.036] | 0.549 | 0.58 |
| **Salience** | Timepoint | -0.0102 | [-0.033, 0.013] | -0.867 | 0.39 |
| **Salience** | Intervention | -0.0001 | [-0.050, 0.049] | -0.003 | 1.00 |
| **Salience** | Timepoint x Intervention | 0.0136 | [-0.020, 0.047] | 0.802 | 0.42 |
| **DMN** | Timepoint | -0.0188 | [-0.068, 0.030] | -0.754 | 0.45 |
| **DMN** | Intervention | -0.0006 | [-0.074, 0.073] | -0.017 | 0.99 |
| **DMN** | Timepoint x Intervention | 0.0098 | [-0.060, 0.080] | 0.275 | 0.78 |
| **CEN-Salience** | Timepoint | 0.0083 | [-0.007, 0.023] | 1.082 | 0.28 |
| **CEN-Salience** | Intervention | -0.0070 | [-0.038, 0.024] | -0.445 | 0.66 |
| **CEN-Salience** | Timepoint x Intervention | -0.0101 | [-0.032, 0.012] | -0.911 | 0.36 |
| **CEN-DMN** | Timepoint | -0.0140 | [-0.036, 0.008] | -1.230 | 0.22 |
| **CEN-DMN** | Intervention | -0.0262 | [-0.062, 0.009] | -1.441 | 0.15 |
| **CEN-DMN** | Timepoint x Intervention | 0.0189 | [-0.013, 0.051] | 1.155 | 0.25 |
| **Salience-DMN** | Timepoint | -0.0104 | [-0.037, 0.016] | -0.768 | 0.44 |
| **Salience-DMN** | Intervention | -0.0352 | [-0.083, 0.012] | -1.453 | 0.15 |
| **Salience-DMN** | Timepoint x Intervention | 0.0001 | [-0.038, 0.038] | 0.005 | 1.00 |

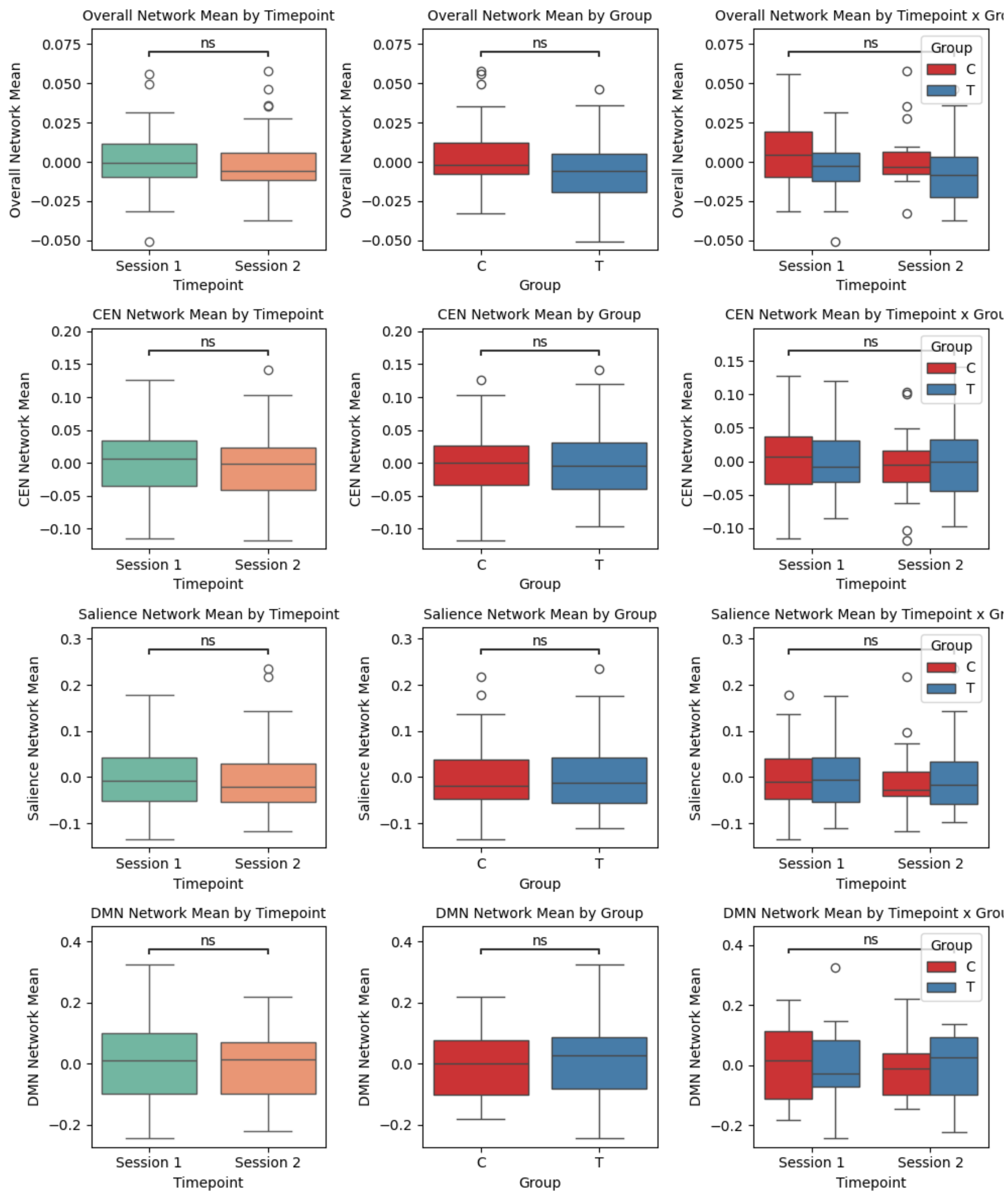


**Supplemental Figure 1 - Boxplot comparisons showing the effects of timepoint, intervention group, and their interaction on within network mean connectivity values.** Column 1 displays differences between baseline (Session 1) and follow-up (Session 2) timepoints across all participants. Column 2 shows differences between control (C) and treatment (T) intervention groups across both sessions. Column 3 illustrates the interaction between timepoint and intervention group, comparing baseline controls to follow-up treatment participants. Statistical significance is derived from mixed-effects linear models accounting for repeated measures within subjects, no effects were significant. Box plots show median (center line), interquartile range (box), and outliers (points beyond whiskers).

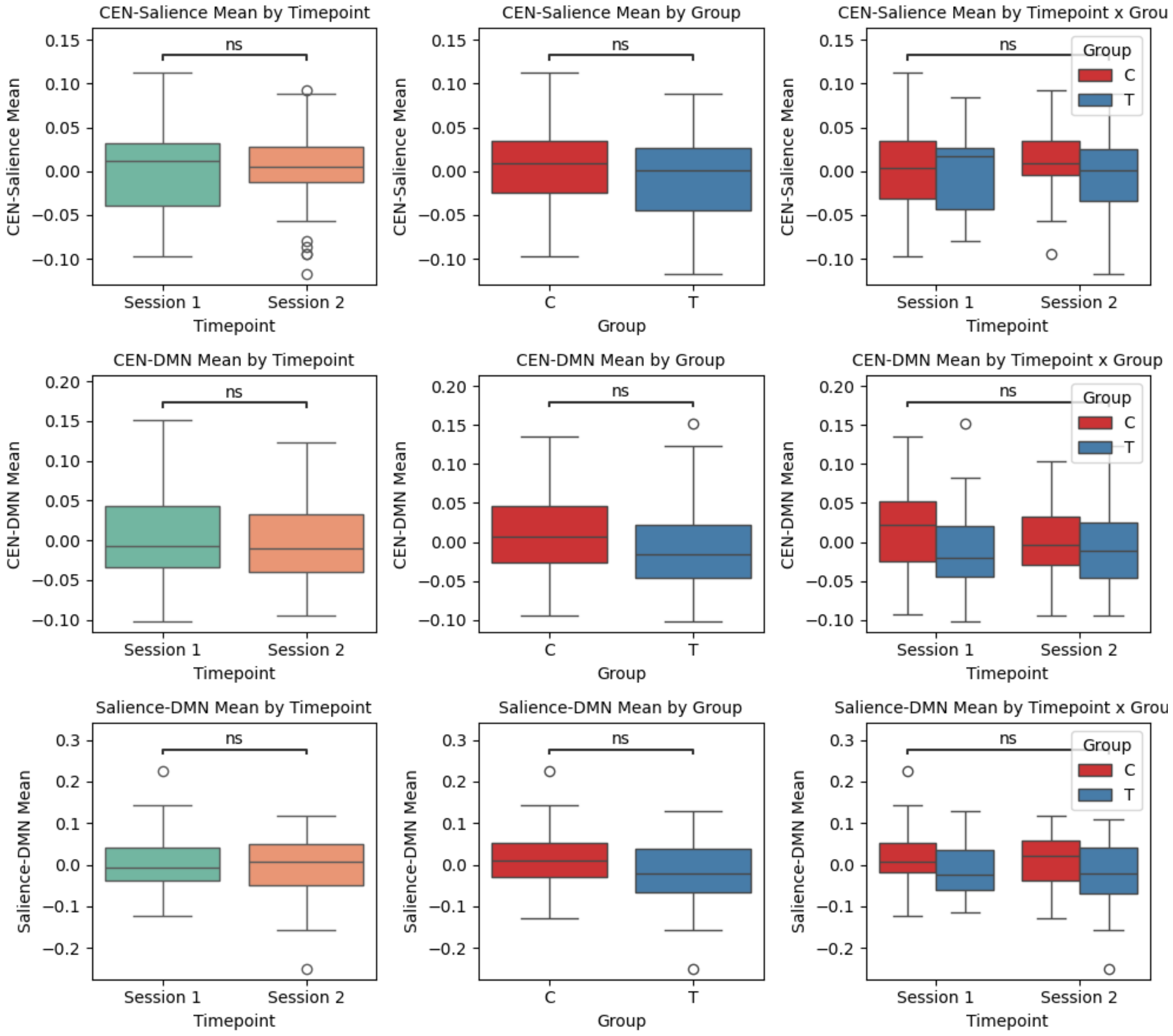


**Supplemental Figure 2 - Boxplot comparisons showing the effects of timepoint, intervention group, and their interaction on between network mean connectivity values.** Column 1 displays differences between baseline (Session 1) and follow-up (Session 2) timepoints across all participants. Column 2 shows differences between control (C) and treatment (T) intervention groups across both sessions. Column 3 illustrates the interaction between timepoint and intervention group, comparing baseline controls to follow-up treatment participants. Statistical significance is derived from mixed-effects linear models accounting for repeated measures within subjects, no effects were significant. Box plots show median (center line), interquartile range (box), and outliers (points beyond whiskers).
