## Supplementary material for "Resting-State Functional Connectivity and Cognitive Performance in Long COVID: A Randomized Controlled Trial": RCT Protocol

**Assessing the Effectiveness of Cognitive Rehabilitation for Post-COVID-19 Cognitive Impairment: A Pilot Study**

### Background

Studies have shown that SARS-CoV-2 infection is associated with post-acute cognitive impairment (CI), which may portend significant consequences for patients' function and quality of life.[1-5] Commonly referred to as "brain fog" and subsumed under "long-COVID," post-COVID-19 CI can persist several months after acute infection and is often described as difficulty concentrating, confusion, and memory difficulties that may result in functional disability.[6,7] Our preliminary data and others' suggest that impairments in executive functions (EF) may be the most prominent deficit in post-COVID-19 CI across the spectrum of acute COVID-19 severity.[8] Despite the significant impact of CI in this relatively young population, there currently are no available interventions with demonstrated efficacy. As one of the most common Long-COVID symptoms, it is imperative that cognitive interventions be developed to help patients return to their baseline level of functioning.

Meta-analyses have shown that cognitive rehabilitation (CR) interventions improve cognitive, emotional, and functional outcomes in various clinical and healthy aging populations.[9,10] CR is linked to the concept of neuroplasticity whereby, through practice, the brain can create new neural connections, or strengthen existing ones.[11] This leads to reinforcement of the trained cognitive capacity (i.e., near effects), and often to related abilities as well (i.e., far effects). While typically tailored to patient-specific cognitive deficits, CR specifically for attention and executive functioning has shown the greatest efficacy in terms of cognitive improvement across clinical populations.[12,13] Accordingly, neuroimaging studies have consistently shown structural and functional changes of executive dysfunction after CR in participants with brain injury (e.g., stroke, traumatic brain injury, chemotherapy-related CI).[14,15] Given accumulating evidence of COVID-19's potential to injure the brain,[16] together with objective data of post-COVID-19 CI across studies, it is likely that post-COVID-19 patients with CI could derive significant benefit from CR. However, there is no empirical data showing the benefits of CR in this population.

Research on the neurobiological mechanisms that underlie CR-induced cognitive changes are only beginning to be understood. As the field evolves from questions of efficacy into mechanisms of action, neuroimaging may help to understand links between cognitive change (e.g., improvement on a clinical measure) and actual neurologic change. Resting state functional magnetic resonance imaging (rsfMRI) can provide valuable information about the intrinsic connectivity of specific cognition-related brain networks, allowing us to assess relationships between impairments in brain networks and cognitive performance. The central executive network (CEN),[17] a frontoparietal brain network underlying EF, may be particularly implicated in post-COVID-19 CI. As such, examination of CEN connectivity leveraging rsfMRI could provide insight into the mechanisms underlying the impact of CR in post-COVID-19 CI. An increased understanding of the capacity of CR to induce functional neurologic change post-COVID-19 has the potential to maximize the clinical relevance of CR interventions by allowing researchers to optimize CR programs and potentially predict treatment responses.

The goal of this pilot randomized controlled trial is to investigate the feasibility and efficacy of CR for patients with post-COVID-19 CI. Additionally, we will obtain preliminary data, using rsfMRI, regarding the potential underlying connectivity networks, mediating the effect of CR on cognitive improvements.

Our Specific Aims are to:

**Aim 1: Determine the feasibility and preliminary effectiveness of an adaptation of a CR protocol for patients with post-COVID-19 CI**

**Aim 2: Examine whether changes in CEN connectivity mediate the effect of CR on cognition in patients with post-COVID-19 CI**

**Overall Approach:** This pilot study will assess the feasibility and provide preliminary data about the efficacy and associated neural changes of a 12-week CR intervention for patients with post-COVID-19 CI. The CR intervention will be conducted virtually and will specifically target post-COVID-19 executive dysfunction. Cognitive assessment and rsfMRI will occur prior to and following (within two weeks) the CR intervention.

The attention control group will receive a parallel control intervention as described in section 1.3, and control participants will be offered a delayed CR intervention after completing the study.

**Research Design and Methods:** We will randomize a total of 60 patients with post-COVID-19 CI to either CR vs. an attention control.

**1.1 Settings and Participants**: Participants will be recruited from the Mount Sinai Health System's (MSHS) Post-COVID-19 Registry or MSHS Center for Post-COVID-19 care. Inclusion criteria are: 1) adults ≥18 years of age, 2) have a confirmed SARS-CoV-2 positive test or are serum antibody positive (against spike protein if unvaccinated or N-capsid peptide if vaccinated), 3) speak English, 4) have objective mild to moderate CI, which will be defined as impairment in EF (a z-score >1.5 standard deviations below the normative mean) and in at least one other cognitive domain, and 5) endorse comfort and familiarity with technology. Exclusion criteria: 1) no method of contact, 2) major neurocognitive disorder, as defined by a score <17 on the Montreal Cognitive Assessment (MoCA),[18] 3) history of pre-COVID-19 neurologic disease (e.g., stroke), 4) history of severe head injury (as defined by loss of consciousness >30 minutes), 5) ongoing substance use disorder (Mini International Neuropsychiatric Interview Version 7.0.2 [MINI] or Alcohol Use Disorders Test-Consumption [AUDIT-C] >8), 6) unstable medical, neurologic, or psychiatric conditions precluding participation in research activities, and 7) contraindication for MRI (e.g., metallic/electronic implants). We will ensure adequate representation of women and minorities that reflects the population of COVID-19 survivors in our region.

**1.2 Data Collection**: Participants will undergo an in-person baseline interview conducted by trained research coordinators at the Center for Advanced Medicine (CAM) building. The baseline interview will consist of questionnaires assessing sociodemographic characteristics, pre-COVID-19 health status, and acute COVID-19 presentation and treatment. Trained research coordinators will administer cognitive functioning assessments at baseline, within two weeks of beginning the intervention. rsfMRI scanning will also occur in the same visit. Cognitive assessments and rsfMRI scanning will then be repeated within two weeks of intervention completion. Participants will be compensated $50 at the baseline visit and $75 for the post-intervention visit. Data will be collected in REDCap, verified, cleaned, recoded to compute scored measures, and saved under password and encryption protection in the MSHS cloud.

**1.3 Intervention procedures**: After baseline assessment, eligible participants will be randomly allocated to the intervention or the attention control group using an optimization algorithm to minimize imbalances in age, sex or baseline CI severity between groups (Figure 1).

Figure 1. Study Design

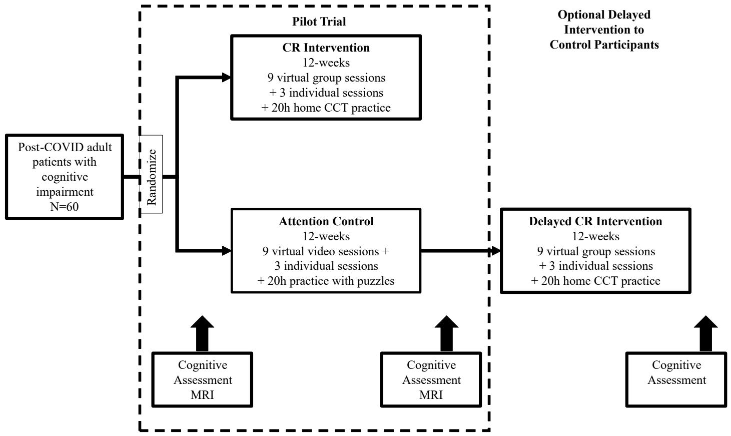


Active Group: Participants in the active arm will receive an adaptation of the Goal-Oriented Attentional Self-Regulation (GOALS)[19] intervention in a virtual group format. The GOALS intervention is a manualized treatment protocol initially developed for individuals with chronic sequela of traumatic brain injury with demonstrated efficacy in improving cognitive functioning. GOALS is an intervention that typically emphasizes two key components: 1) attention regulation training (i.e., redirection of attention to goal-relevant processes and the filtering of non-relevant ''noise,'' especially in the context of distractions) and 2) application of the goal-oriented attentional self-regulation skills to the identification, selection, and execution of self-generated complex goals. As part of the adaptation for patients with post-COVID-19, we will implement virtual sessions, and we will incorporate two new components that are salient for the population with post-COVID-19 symptoms: a) fatigue management training, and b) computerized cognitive training (CCT) using BrainHQ (Posit Science).

The first component above emphasizes principles of applied mindfulness-based attention regulation to redirect cognitive processes towards task-relevant activities even when distracted. This requires identifying the current primary task, separating information into relevant and non-relevant categories, and working to selectively maintain relevant information while letting go of non-relevant information. Participants are trained in applying a meta-cognitive strategy (‘‘Stop-Relax-Refocus’’) to stop activity when distracted, anxious, and/or overwhelmed; relax; and then re-focus attention on the current primary goal. They are taught to actively apply goal-directed attention regulation skills to a range of situations, from simple information processing tasks to complex multi-step problems and challenging low-structure situations occurring in their own lives. Training via in-class exercises and homework is applied to progressively more challenging situations, including holding increasing amounts of information in mind, culminating with maintaining information while exposed to distractors. Homework includes practice in maintaining goal-direction during challenging real-life situations identified by participants. At the beginning of each session, participants discuss their experiences completing the homework with the rest of the group. The second component of GOALS training involves asking participants to apply the above skills to identify, select, and execute self-generated and personally relevant feasible functional goals (e.g., planning a vacation, completing a work assignment). Participants are then trained in applying the goal management strategies on the functional task(s) of their choice. The step-wise meta-cognitive goal management strategies trained were modified from the Goal Management Training and Problem Solving Therapy protocols. The main objective is to allow extensive practice and application of skills, thereby linking the attentional regulation directly to goal attainment efforts, which can improve attention and EF. The third component of fatigue management training will address the cognitive fatigue that is commonly reported post-COVID-19. The goal of this component is to teach participants compensatory strategies for managing cognitive fatigue, including “brain breaks”, planning ahead, and avoiding overstimulation. It also involves behavioral activation with a focus on restoring functionality. The final component involving CCT is a structured, computerized brain-training program, which trains individuals with a schedule of 23 pre-selected exercises in 20- to 25-minute sessions (6-10 exercises per session). All exercises will target the speed and accuracy of neural information processing, requiring attentional focus to perform correctly. The content is delivered based on algorithms that ensure an appropriate level of challenge, which continuously adjusts across five levels of activity, depending on how the participant is performing that session. Although all participants will receive comparable training with equal emphasis among domains, the exact sequence of exercises depends on each participants’ ability level. The session-by-session content from the GOALS intervention is depicted in **Table 1**. **Attention regulation**

**Table 1. Session content**

| **Session** | | **Content** |
|---|---|---|
| Group #1 | **Attention Regulation Training<br>Applied Goal Management Training<br>Fatigue Management Training** | Introduction and overview |
| **Individual #1** | | Individualized assessment and strategy-building |
| Group #2 | | Progressive information maintenance and mindfulness exercises |
| Group #3 | | Goal selection: options for group and individual projects |
| Group #4 | | Breaking down projects into sub-tasks, creating timeline. Apply to group and individual projects |
| Group #5 | | Execution and dealing with procrastination |
| **Individual #2** | | Individualized progress monitoring |
| Group #6 | | Staying on tasks, error correction, and adjustments |
| Group #7 | | Project progress review and adjustments |
| Group #8 | | Individual project presentation |
| Group #9 | | Group project presentation, discussion of maintenance, and graduation celebration |
| **Individual #3** | | Individual wrap-up |

**training**, **applied goal management training**, and **fatigue management training** will be applied across all sessions.

Participants will receive nine 1.5-h sessions of **virtual** (via Zoom platform) group-based training, three individual **virtual** 1-h training sessions, and approximately 20 h of home practice and CCT over 12 weeks (Figure 2). The virtual small group format will include two to five participants and one facilitator per group. The three individual sessions are distributed at the beginning, middle, and towards the end of the training program. Individual Session 1 occurs after Group Session 1. It covers orientation to the training program, including orientation to and clarification of the Participant Handbook. Individual Session 2 occurs after Group Session 5 and involves review and implementation of training strategies. Individual Session 3 occurs after Group Session 9 and covers review of learned strategies, implementation of training strategies in daily life, and review of execution, obstacles, and adjustments required for individual projects.

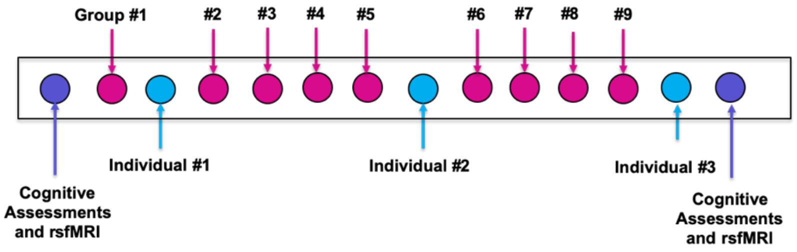

**Figure 2. Intervention sessions overview**

Attention Control Group: The control group format will be twelve 1.5 or 1 h **virtual** sessions over 12 weeks, in addition to 20 h of home practice of a computerized control program (e.g., consisting of publicly available computer games such as sudoku and crossword puzzles). The active control intervention will control for non-specific effects inherent to supervised CR. Participants will view seven short National Geographic videos per group session on a computer and answer multiple-choice questions immediately after each presentation. Once these participants complete the 12 weeks of the control intervention and undergo the post-intervention assessments (see below), they will be offered the 12 weeks active intervention to ensure that all study participants have the potential of benefitting from CR.

**1.4 Study Outcomes**:

1) Cognitive Functioning**:** We will include a cognitive battery measuring the following domains: attention, working memory, processing speed, EF, learning and memory, and language. EF will be the primary outcome and will be defined as impairment on at least one of the two EF measures described below. We will analyze other cognitive domains (e.g., attention, working memory, processing speed, and language) as secondary outcomes. We will randomly counterbalance alternate forms for applicable measures to reduce practice effects (PE). (a) EF: TMT-Part B[20,21] is a measure of set- shifting and cognitive flexibility, where participants connect the dots on a page scattered with numbers and letters in alternating sequence (reliability 0.60-0.90). The Wisconsin Card Sorting Test-64 (WCST-64)[21,22] is a measure assessing novel problem solving and abstract thinking. (b) Attention and working memory: In the Number Span from the National Alzheimer's Coordinating Center (NACC) battery[20] participants must recall increasingly longer lists of numbers, first forward, then backward. The Paced Auditory Serial Addition Task (PASAT)[23] is a measure of sustained attention and information processing that requiring examinees to be vigilant to stimuli (numbers) quickly presented through an audio recording. (c) Processing speed: The Symbol Digit Modalities Test (SDMT)[24] requires an individual to substitute digits for abstract symbols using a reference key as quickly as possible. The Trail Making Test (TMT)-Part A[20,21] requires participants to connect numbered dots in order as quickly as possible. (d) Learning and Memory: The Hopkins Verbal Learning Test- Revised (HVLT-R),[25-28] is a list learning task that assesses learning (immediate recall) and memory (delayed recall) and recognition discrimination, the latter of which will also be used as an embedded measure of performance validity, which has been empirically validated.[29,30] The Brief Visuospatial Memory Test-Revised (BVMT-R)[27,31] requires reproduction of two-dimensional geometric figures from memory, assessing visuospatial learning (immediate recall) and memory (delayed recall) and recognition. (e) Language: The NACC battery's verbal phonemic and category fluency tests[20] requires participants to name as many words as they can beginning with a particular letter in 60 seconds, and then as many animals as they can, respectively. The Multilingual Naming Test (MINT)[20] will assess confrontation naming. We will also assess reading ability as a proxy for premorbid intellectual ability and quality of education using a culturally-sensitive measure with high test-retest reliability. (f) Premorbid intellectual ability: The Wide

Range Achievement Test, 4th ed. (WRAT-4)[32] reading subtest is among the most valid estimates of quality of education among multiethnic, English-speaking samples, with high test-retest reliability. It requires participants to read a list of words outloud; scoring is based on correct pronunciation of a word. (g) Subjective cognitive complaints: The Cognitive Failures Questionnaire[33] is a self-reported inventory measuring subjective cognitive complaints on a Likert-type scale. Scoring of Cognitive Measures: All scores will be transformed into *z*-scores using published normative data. In exploratory analyses, we will also create an overall Attention and EF score by averaging total scores from attention and EF measures. This composite *z*-score will range from -3 to 3, with higher scores indicating better performance.

2) Feasibility Outcomes: We will evaluate four feasibility metrics based on an established framework.[34,35] 1) We will measure acceptability using 7 Likert-rated items adopted from Manne et al.[36] (scores will be coded as: Low: ≤3; Moderate: 3-3.9; Good: ≥4). We will also use open ended questions to solicit feedback from participants and facilitators regarding how the intervention might be changed. 2) Practicality, or ability to carry out the intervention, will be measured using two indicators: enrollment yield (# enrolled/approached) of >70% and dropout <20%; and open-ended questions to staff regarding challenges conducting intervention (e.g., difficulty enrolling, reaching, engaging participants; scores will be coded as: Low: <50%; Moderate: 50-70%; Good: >70%). 3) Adaptation, or how well the modified intervention performs in a virtual format with post-COVID-19 patients, will be measured through participant and facilitator feedback regarding intervention format and attendance to scheduled sessions. 4) Outcome Expectancy: We will evaluate participants' beliefs about how likely they are to benefit from the intervention using the Treatment Credibility and Expectancy Questionnaire. The scale assesses both expectancy and credibility.[37]

**1.5 Neuroimaging Outcomes**: Our primary neuroimaging outcome will be resting state functional connectivity (rsFC) within the central executive network (CEN), a network underlying EF, the most prominent deficit post-COVID-19.[8] In **secondary analyses**, we will examine rsFC between the CEN and two other large-scale networks involved in cognitive functioning: 1) rsFC between CEN and the default mode network (DMN), which has been linked to memory impairment,[38] 2) rsFC between CEN and the salience network (SN), and 3) rsFC within and between DMN and SN. In **exploratory analyses**, we will quantify white matter hyperintensities (whole brain T2/FLAIR), coded according to the Fazekas scale.[39]

**1.6 Imaging Acquisition:** Participants will undergo 3T MRI scanning at the BioMedical Engineering and Imaging Institute. A standard neuroimaging approach will be utilized. Structural imaging allows for visualization and analysis of brain anatomy, and can be useful to detect damage and abnormalities, as well as for quantifying volume or thickness of a given region.[40] rsfMRI, our primary measure, can identify underlying neural processes by evaluating altered interactions between different brain regions, known as networks.[41] This approach can be used to examine if the brain's functional organization is altered due to a particular condition or following a particular treatment or intervention. Scans will be acquired on a Siemens 3T Skyra scanner using a 32-channel phased array head coil. Total scan time will be one hour. Our protocol has been optimized across many studies[42] and functions robustly on our 3T scanner. All MRIs will be reviewed by a neuroradiologist who will report on incidental findings.

We will conduct seed-based correlation analysis (SCA)[43] in AFNI[44] using established methods[43,45-48] For each voxel, raw time-series data are de-meaned and converted to percentage-signal-change scores to reduce variability between participants. The mean timeseries of activity across voxels within each CEN seed will be used as a temporal predictor in whole-brain regression analyses; the resulting correlation coefficients will be converted to normally distributed z-values. For each CEN seed, rsFC is quantified as the average correlation of its time series to that of all other regions of interest (ROI) within the network. Next, the z-scores of all possible connections are averaged to compute a total network value reflecting the connectivity of all possible ROIs within the network. CEN seed ROIs are defined using a 5-mm radius sphere centered at coordinates for 10 CEN nodes (**Table 2**)[46] identified via independent components analysis and included in a publicly

**Table 2. Regions of Interest for the CEN and coordinates**

| Regions of Interest (ROIs) | Coordinates (x, y, z) |
|---|---|
| 1. Left inferior parietal lobule | -42, -63, 46 |
| 2. Left middle frontal gyrus | -32, 23, 49 |
| 3. Left middle frontal gyrus | -40, 48, -01 |
| 4. Left middle temporal gyrus | -59, 42, 12 |
| 5. Left medial frontal gyrus | -07, 34, 43 |
| 6. Right middle frontal gyrus | 38, 26, 42 |
| 7. Right inferior parietal lobule | 48, -54, 47 |
| 8. Right middle frontal gyrus | 38, 54, 01 |
| 9. Right Caudate | 13, 02, 14 |
| 10. Right medial frontal gyrus | 06, 37, 46 |

available atlas of resting state networks.[58] Hence, using these data and prior methods[46-48,58] our primary neuroimaging outcome will be a single *z*-score reflecting overall rsFC within the CEN. We will use a similar approach to quantify inter- and intra-network connectivity of the DMN (utilizing ROIs listed in Anderson et al[59]), and SN (utilizing ROIs listed in Nusslock et al[47]).

| Table 3. Summary of Study Measures | | | |
|---|---|---|---|
| **Cognitive Domains** | **Measures** | **Time point** | |
| | | **BL** | **Post** |
| Executive Functioning | Trail Making Test- Part B[20,21]<br>Wisconsin Card Sorting Test- 64[21,22] | x<br>x | x<br>x |
| Attention | Number Span Forward[20]<br>Paced Auditory Serial Addition Task (PASAT)[23] | x | x |
| Working Memory | Number Span Backwards[20] | x | x |
| Processing Speed | Trail Making Test- Part A[20,21]<br>Symbol Digit Modalities Test (SDMT)[24] | x | x |
| Learning | Hopkins Verbal Learning Test- Revised[25-28] (Immediate)<br>Brief Visuospatial Memory Test- Revised[56] (Immediate) | x<br>x | x<br>x |
| Memory | Hopkins Verbal Learning Test- Revised[25-28] (Delayed)<br>Brief Visuospatial Memory Test- Revised[56] (Delayed) | x<br>x | x<br>x |
| Language | Verbal Phonemic and Category Fluency[20]<br>Multilingual Naming Test (MINT)[20] | x | x |
| Premorbid Intellectual Ability | Wide Range Achievement Test-4 (English);[32] | x | |
| Subjective cognitive complaints | Cognitive Failures Questionnaire[33] | x | x |
| **Feasibility Outcomes** | | | |
| Acceptability | 7 Likert-rated items adopted from Manne et al.[36] | | x |
| Practicality | Enrollment yield and open-ended questions | | x |
| Adaptation | Participant and facilitator feedback | | x |
| Outcome Expectancy | Treatment Credibility and Expectancy Questionnaire[37] | | x |
| **Covariates** | | | |
| Sociodemographics | Age, sex, race/ethnicity, education, etc.[49] | x | |
| Comorbidities | Specific Comorbidities, Charlson Comorbidity Index[50-52] | x | |
| Depression | Patient Health Questionnaire-8[53] | x | |
| Anxiety | Generalized Anxiety Disorder[54] | x | |
| PTSD | PTSD Checklist, DSM-5[55,56] | x | |
| Fatigue | PROMIS-29 v2.1 Fatigue Subscale[57] | x | x |
| IADLs | Functional Activities Questionnaire[87] | x | x |
| COVID-19 Vaccination | Self-report | x | |
| COVID-19 Variant | Date of acute COVID-19 episode | x | |

**1.7 Covariates:** 1) <u>Sociodemographic variables</u>: age, sex, race, ethnicity, country of birth, native language, education, marital/partner status, income, employment, and smoking history.[49] 2) <u>Pre-COVID-19 health</u>: self- reported data using National Health Interview Survey[49] questionnaires on pre-COVID-19 comorbidities known to impact cognition[60] including hypertension, hyperlipidemia, cardiovascular disease, diabetes chronic obstructive pulmonary disease, depression, memory problems, and obesity (based on body mass index).[61] Global burden of comorbidities will be assessed using the Charlson Comorbidities Index.[51,52] 3) <u>Post-COVID-19 health</u>: data on current symptoms that may impact CI will also be assessed, including fatigue (PROMIS-29 v2.1),[57] instrumental activities of daily living (IADLs), and psychological morbidity (i.e., depression symptoms [PHQ-8],[53] anxiety symptoms [GAD-7],[54] PTSD symptoms [PCL-5]).[55,56] 4) <u>Vaccination status</u>: data on number of doses and type of vaccine and timing in relationship with COVID-19 (pre- vs. post). 5) <u>COVID-19 Variants</u>: date of acute COVID-19 will be a proxy for COVID-19 variants (e.g., Alpha, Delta, Omicron).

<u>Data Analysis</u>: First, we will compare the baseline characteristics of participants randomized to the active intervention vs. control group using a Student's T-test or chi-square test, as appropriate. We will use a one sided Student's T-test (alpha level 0.1) to assess the futility of the intervention by comparing pre vs. post intervention

changes in EF and other cognitive scores in the intervention vs. control group (see Sample Size justification). Similar analyses will be used to compare changes in CEN scores according to treatment arm. Other cognitive measures and connectivity networks will be compared in secondary exploratory analyses. We will adjust or critical value for secondary analyses to control our false discovery rate. Feasibility: will be examined using benchmarks as described in section 1.4. Acceptability will be assessed using mean scores from Likert questions (Low: ≤3; Moderate: 3-3.9; Good: ≥4). For practicality, we will calculate enrollment yield and dropout rates; and compare the 95% upper bound for observed rates to targeted rates (Low: <50%; Moderate: 50-70%; Good: >70%). To analyze the open-ended questions, we will use a general inductive approach. In exploratory analyses, we will use structural equation modeling to test if changes in CEN between study arms mediate the effect of the intervention.
